# Patient-reported predictors of post-discharge mortality after cardiac hospitalization

**DOI:** 10.1101/2023.10.02.23296460

**Authors:** Devika Nair, Jonathan S. Schildcrout, Yaping Shi, Ricardo Trochez, Sam Nwosu, Susan P. Bell, Amanda S. Mixon, Sarah A. Welch, Kathryn Goggins, Justin M. Bachmann, Eduard E. Vasilevskis, Kerri L. Cavanaugh, Russell L. Rothman, Sunil B. Kripalani

## Abstract

**Background:** Adults hospitalized for cardiovascular events are at high risk for post-discharge mortality. Hospital-based screening of health-related psychosocial risk factors is now prioritized by the Joint Commission and the National Quality Forum to achieve equitable, high-quality care. We tested our hypothesis that key patient-reported psychosocial and behavioral measures could predict post-hospitalization mortality in a cohort of adults hospitalized for a cardiovascular event.

**Methods:** This was a prospective cohort of adults hospitalized at Vanderbilt University Medical Center. Validated patient-reported measures of health literacy, social support, disease self-management, and socioeconomic status were used as predictors of interest. Cox survival analyses of mortality were conducted over a median 3.5-year follow-up (range: 1.25 – 5.5 years).

**Results:** Among 2,977 adults, 1,874 (63%) were hospitalized for acute coronary syndrome and 1,103 (37%) were hospitalized for acute decompensated heart failure; 60% were male; and the mean age was 53 years. After adjusting for demographic, clinical, and other psychosocial factors, mortality risk was greatest among patients who reported being unable to work due to disability (Hazard Ratio (HR) 2.36, 95% Confidence Interval (CI): 1.73-3.21), who were retired (HR 2.14, 95% CI 1.60-2.87), and who reported unemployment (HR 1.99, 95% CI 1.30-3.06) as compared to those who were employed. Patient-reported measures of disease self-management, perceived health competence and exercise frequency, were also associated with mortality risk after full covariate adjustment (HR 0.86, 95% CI 0.73-1.00 per four-point increase), (HR 0.86, 95% CI 0.77-0.96 per three-day change), respectively.

**Conclusions:** Patient-reported measures of employment status independently predict post-discharge mortality after a cardiac hospitalization. Measure of disease self-management also have prognostic modest utility. Hospital-based screening of psychosocial risk is increasingly prioritized in legislative policy. Incorporating brief, valid measures of employment status and disease self-management factors may help target patients for psychosocial, financial, and rehabilitative resources during post-discharge transitions of care.

## Introduction

Mortality after hospitalization for acute coronary events and acute decompensated heart failure is high, and ranges from 12-20%.^1-2^ Valid, feasible methods to identify patients at greatest risk for mortality after discharge are necessary to improve care transitions and optimally target follow-up support programs for these vulnerable groups. The contribution of medical factors including primary disease status and comorbidities on mortality risk have been long-established, as has the general importance of social and behavioral factors.^3^ However, only recently have hospitals and health systems begun to collect data on psychosocial and behavioral risk factors for health outcomes to inform patient care.^4-5^ New regulations from The Joint Commission now require hospitals to collect data on health-related psychosocial risk factors and will increase the availability and potential uses of these data.^6^

Patient-reported psychosocial and behavioral measures have the potential to identify those at greatest risk of post-discharge adverse outcomes, including mortality.^7^ Psychosocial measures include socioeconomic status, perceived social support, subjective and functional health literacy and numeracy; behavioral measures include alcohol use and aspects of disease self-management such as perceived health competence and medication adherence. These measures are known to add significant prognostic value to predictive models that estimate mortality risk among individuals living with a malignancy,^7^ but their prognostic utility in vulnerable populations after an acute hospitalization is less well-studied. This gap in knowledge limits the health system’s ability to optimally target resources to improve transitions of care after an acute hospitalization.

Informed by a thorough literature review of psychosocial and behavioral measures that predict mortality,^8-20^ we tested the hypothesis that patient-reported health literacy, social support, disease self-management, and socioeconomic status would independently predict mortality after hospitalization for acute coronary syndrome or acute decompensated heart failure, after adjustment for demographic and clinical factors.

## Methods

### Study design and sample

This analysis is part of the National Heart Lung and Blood Institute-funded Vanderbilt Inpatient Cohort Study (VICS), aimed to identify novel, modifiable factors that predict adverse outcomes during the post-hospitalization period.^21^ Briefly, VICS was a prospective cohort that included 2,977 adults hospitalized for acute decompensated heart failure or acute coronary syndromes at Vanderbilt University Hospital in Nashville, Tennessee between 2011 and 2015. The cohort is unique in its administration of a wide range of validated patient-reported measures at cohort enrollment. Eligibility was determined from medical record review conducted by a physician and research staff. Criteria for exclusion were severe cognitive impairment, unstable psychiatric illness, inability to communicate in English, current hospice status, and unstable contact information for follow-up. For this analysis, we excluded patients who died during the index hospitalization and therefore did not enter the post-discharge follow-up period. The study was approved by Vanderbilt’s Institutional Review Board, and written informed consent was obtained from enrollees.

Demographic factors collected included age, gender, years of maximum educational attainment, household income, self-reported race, and marital status. Number of hospitalizations during the prior 12 months and a validated score of total clinical comorbidity burden (the Elixhauser Comorbidity Index)^22^ were also obtained. The Elixhauser Comorbidity Index includes whether a participant has congestive heart failure, cardiac arrythmias, cardiac valvular disease, pulmonary hypertension, peripheral vascular disease, hypertension, paralysis, neurodegerative disordres, chronic obstructive pulmonary disease, diabetes, hypothyroidism, kidney disease, liver disease, peptic ulcer disease, acquired immunodeficiency symdrome, lymphoma, metastatic or solid malignancies, collagen vascular disease, coagulopathy, obesity, weight loss, fluid and electrolyte disorders, anemia, evidence of alcohol or illicit substance abuse, depression, or psychosis. Cause of cardiovascular hospitalization (acute coronary syndrome vs. acute decompensated heart failure) were ascertained from the medical record.

### Exposures/predictors of interest

We identified validated, patient-reported measures of both functional and subjective health literacy, subjective numeracy, social support, and health behaviors as worthy of further investigation for prognostic utility in our sample. Given the known significant associations between self-efficacy and health outcomes, including mortality, across multiple chronic conditions^23-24^ we also tested whether a validated measure of health self-efficacy, perceived health competence, was associated with mortality in our sample. Given that health behaviors and emotion management are encompassed by a construct known in the literature as disease self-management,^25^ we grouped patients’ reports of health behaviors, coping, and perceived health competence into a ‘Disease Self-Management’ domain.

Functional health literacy was measured using the short form of the Test of Functional Health Literacy in Adults (sTOFHLA; scores 0-36 with higher scores indicating greater ability to interpret health-related text). This test categorizes functional health literacy as inadequate (Score 0-16), marginal (score 17-22), or adequate (scores 23-36).^26^ Subjective health literacy was assessed using the three-item Brief Health Literacy Screen (BHLS; scores 3-15 with higher scores indicating higher confidence in interpreting written and spoken health-related material).^27^ Numeracy was measured using a three-item version of the Subjective Numeracy Scale (SNS; scores 1-6 with higher scores indicating greater comfort with interpreting numbers).^28^

Social support was assessed using multiple measures, each of which identified a different aspect of social support. These included items from the Health and Retirement Survey and items from the Midlife Development in the United States (MIDUS) survey that measured frequency of contact and level of support from friends, family, and neighbors.^29-30^ We also added the Enhancing Recovery in Coronary Heart Disease Emotional Social Support Instrument (ESSI; scores 8-34 with higher scores indicating greater perceived emotional support).^31^

Disease self-management was assessed using patients’ frequency in engaging in health behaviors and ability to manage emotional stressors associated with living with a health condition.^25^ Perceived health competence was assessed using a validated, two item abbreviation of the original, eight-item Perceived Health Competence Scale, a measure of individuals’ self-confidence in managing health and achieving health goals (PHCS-2; scores 2-10 with higher scores indicating higher perceived health competence).^20,32^ Resilient coping behaviors, the ability to rebound from or positively adapt to significant stressors, was measured by the validated four-item Brief Resilient Coping Scale (BRCS; scores 4-20 with higher scores indicating a greater ability to cope with stress).^33^ Medication adherence before hospitalization was measured using a seven-item version of the Adherence to Refills and Medicines Scale (ARMS, scores 7-28 with higher scores indicating higher self-reported medication adherence).^34^ Tobacco and alcohol use, dietary habits, and weekly exercise frequency were measured using the Centers for Disease Control Behavioral Risk Factor Surveillance System questions, the Starting the Conversation scale, and the Exercise Vital sign scale, respectively.^35-37^

Socioeconomic status was assessed by self-reported annual household income, highest level of education attained, employment status (employed, not employed for wages, retired, or unemployed due to a disability), and degree of difficulty paying bills.

### Outcome variable

Mortality was ascertained using the Social Security Administration’s Death Master File; information from patient charts recorded after clinical contact with family members; and obituaries. The Death Master File contains over 85 million records since 1936 and is linked to patient records by Social Security Number. The Death Master File provides accurate matches for death (in excess of 90% among American born individuals), but has had considerable omissions, particularly since 2011.^38-39^ We therefore supplemented Death Master File data with clinical records and obituaries. Mortality follow-up was for a minimum of 1.25 years for the last patient enrolled (December 2015), and up to 5.5 years for the initial enrollees (October 2011). Therefore, any variability in followup was not due to loss to followup but due to administrative censoring.

### Statistical analyses

We summarized categorical variables with counts and percentages, and continuous variables with the median (25^th^, 75^th^) percentiles. For the primary analyses, we fitted multivariable Cox proportional hazards regression models to investigate the associations between clinical and patient-reported risk factors and time to death.

We defined six domains of variables: 1) demographics and reason for hospitalization (age, gender, race, and acute coronary syndrome vs acute decompensated heart failure), 2) health literacy and numeracy (BHLS, sTOFHLA, SNS), 3) social support (relationship status, number of close family and/or friends, ESSI), 4) disease self-management (PHCS-2, BRCS, ARMS, alcohol use, tobacco use, exercise, dietary habits), 5) socioeconomic status (income, education, employment, difficulty paying bills) and 6) health status (Elixhauser comorbidity score, hospitalizations in prior 12 months). We performed redundancy analysis to examine the potential for collinearity among all risk factors. The highest coefficients of determination corresponded to the income and age values, with values of 0.59 and 0.50, respectively. All other coefficients of determination were 0.4 or lower, and no factors were removed from consideration.

To examine the association of mortality rates and risk factors or domains of risk factors, we implemented a hierarchical modeling approach. We first fit a Base Model that only included variables in the demographics and diagnosis domain. We then added all variables in each of the five other domains (one domain at a time) to the Base Model to examine their associations while adjusting for the Base Model variables but without adjusting for all other domains. Finally, we fit a full model with all variables from all domains simultaneously.

We reported hazard ratios and 95% confidence intervals corresponding to interquartile range changes in continuous variables. To test the association between each domain and mortality rates we used likelihood ratio tests (LRT) with degrees of freedom equal to the number of parameters estimated for the domain. For the domain-specific models, the LRT compares the Base Model to a model that adds a single domain of variables to the Base Model. For the full model results, the LRT compares the full model to a model that excludes all variables within in a single domain. Additionally, in the full model, to quantify the contribution of each risk factor and domain, we calculated a measure of relative explained variation (REV) by dividing the likelihood ratio Chi-square statistic associated with each variable or domain by the total likelihood ratio Chi-square of the full model.

We summarized the missingness rate of each variable and conducted multiple imputation with chained equation, with five imputated datasets using a predictive mean matching algorithm. We conducted a single global test for the proportional hazards assumption across all variables. Because there was not a clear violation of the proportional hazards assumption (p=0.06), we chose to assume proportional hazards for all variables. We used a two-sided p < 0.05 significance level and conducted all analyses in R version 4.3.0 (R Foundation for Statistical Computing, Vienna, Austria).^40-42^

## Results

Our sample consisted of 2,977 patients who survived to discharge; 1,874 (63%) were hospitalized for acute coronary syndrome and 1,103 (37%) were hospitalized for acute decompensated heart failure. The median age was 61, 60% were men, and 85% identified as white. The median years of education was 13, 22% had an income below $20,000, 35% were retired, and 25% reported being unable to work due to a disability (Table 1).

**Table 1:**
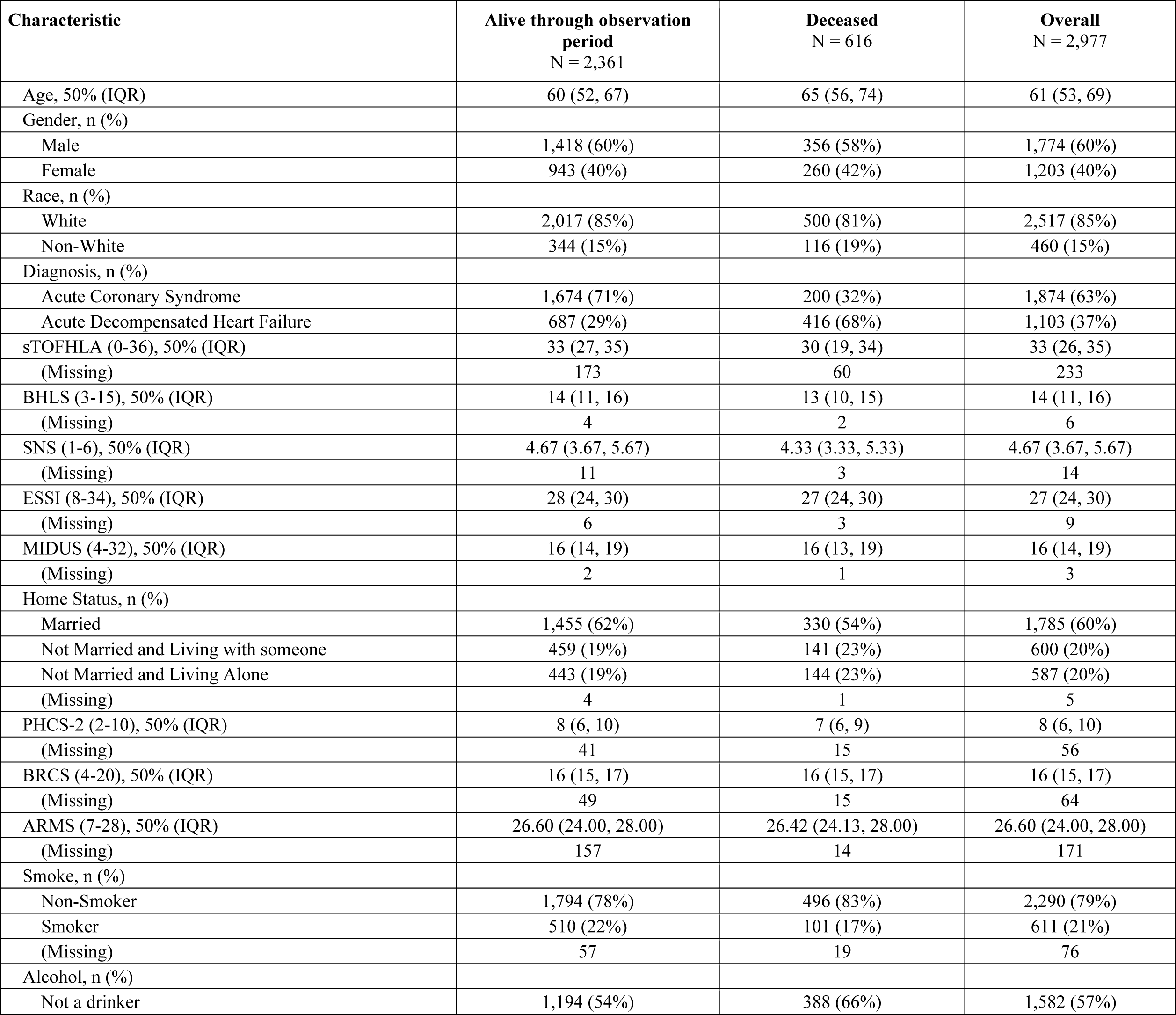
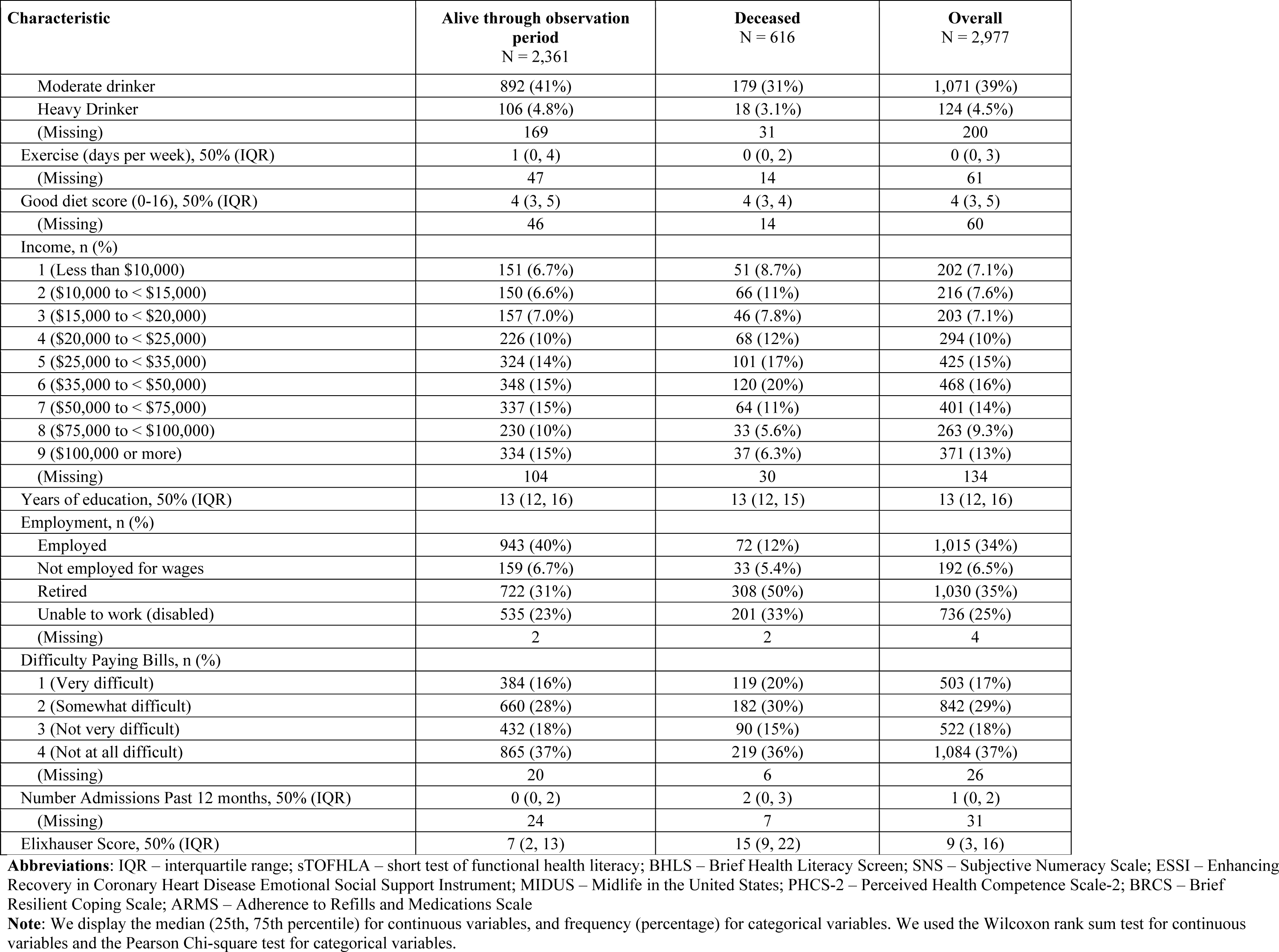
Participant characteristics.

| Characteristic | Alive through observation<br>period<br>N = 2,361 | Deceased<br>N = 616 | Overall<br>N = 2,977 |
| --- | --- | --- | --- |
| Age, 50% (IQR) | 60 (52, 67) | 65 (56, 74) | 61 (53, 69) |
| Gender, n (%) |  |  |  |
| Male | 1,418 (60%) | 356 (58%) | 1,774 (60%) |
| Female | 943 (40%) | 260 (42%) | 1,203 (40%) |
| Race, n (%) |  |  |  |
| White | 2,017 (85%) | 500 (81%) | 2,517 (85%) |
| Non-White | 344 (15%) | 116 (19%) | 460 (15%) |
| Diagnosis, n (%) |  |  |  |
| Acute Coronary Syndrome | 1,674 (71%) | 200 (32%) | 1,874 (63%) |
| Acute Decompensated Heart Failure | 687 (29%) | 416 (68%) | 1,103 (37%) |
| sTOFHLA (0-36), 50% (IQR) | 33 (27, 35) | 30 (19, 34) | 33 (26, 35) |
| (Missing) | 173 | 60 | 233 |
| BHLS (3-15), 50% (IQR) | 14 (11, 16) | 13 (10, 15) | 14 (11, 16) |
| (Missing) | 4 | 2 | 6 |
| SNS (1-6), 50% (IQR) | 4.67 (3.67, 5.67) | 4.33 (3.33, 5.33) | 4.67 (3.67, 5.67) |
| (Missing) | 11 | 3 | 14 |
| ESSI (8-34), 50% (IQR) | 28 (24, 30) | 27 (24, 30) | 27 (24, 30) |
| (Missing) | 6 | 3 | 9 |
| MIDUS (4-32), 50% (IQR) | 16 (14, 19) | 16 (13, 19) | 16 (14, 19) |
| (Missing) | 2 | 1 | 3 |
| Home Status, n (%) |  |  |  |
| Married | 1,455 (62%) | 330 (54%) | 1,785 (60%) |
| Not Married and Living with someone | 459 (19%) | 141 (23%) | 600 (20%) |
| Not Married and Living Alone | 443 (19%) | 144 (23%) | 587 (20%) |
| (Missing) | 4 | 1 | 5 |
| PHCS-2 (2-10), 50% (IQR) | 8 (6, 10) | 7 (6, 9) | 8 (6, 10) |
| (Missing) | 41 | 15 | 56 |
| BRCS (4-20), 50% (IQR) | 16 (15, 17) | 16 (15, 17) | 16 (15, 17) |
| (Missing) | 49 | 15 | 64 |
| ARMS (7-28), 50% (IQR) | 26.60 (24.00, 28.00) | 26.42 (24.13, 28.00) | 26.60 (24.00, 28.00) |
| (Missing) | 157 | 14 | 171 |
| Smoke, n (%) |  |  |  |
| Non-Smoker | 1,794 (78%) | 496 (83%) | 2,290 (79%) |
| Smoker | 510 (22%) | 101 (17%) | 611 (21%) |
| (Missing) | 57 | 19 | 76 |
| Alcohol, n (%) |  |  |  |
| Not a drinker | 1,194 (54%) | 388 (66%) | 1,582 (57%) |
| Moderate drinker | 892 (41%) | 179 (31%) | 1,071 (39%) |
| Heavy Drinker | 106 (4.8%) | 18 (3.1%) | 124 (4.5%) |
| (Missing) | 169 | 31 | 200 |
| Exercise (days per week), 50% (IQR) | 1 (0, 4) | 0 (0, 2) | 0 (0, 3) |
| (Missing) | 47 | 14 | 61 |
| Good diet score (0-16), 50% (IQR) | 4 (3, 5) | 4 (3, 4) | 4 (3, 5) |
| (Missing) | 46 | 14 | 60 |
| Income, n (%) |  |  |  |
| 1 (Less than \$10,000) | 151 (6.7%) | 51 (8.7%) | 202 (7.1%) |
| 2 (\$10,000 to < \$15,000) | 150 (6.6%) | 66 (11%) | 216 (7.6%) |
| 3 (\$15,000 to < \$20,000) | 157 (7.0%) | 46 (7.8%) | 203 (7.1%) |
| 4 (\$20,000 to < \$25,000) | 226 (10%) | 68 (12%) | 294 (10%) |
| 5 (\$25,000 to < \$35,000) | 324 (14%) | 101 (17%) | 425 (15%) |
| 6 (\$35,000 to < \$50,000) | 348 (15%) | 120 (20%) | 468 (16%) |
| 7 (\$50,000 to < \$75,000) | 337 (15%) | 64 (11%) | 401 (14%) |
| 8 (\$75,000 to < \$100,000) | 230 (10%) | 33 (5.6%) | 263 (9.3%) |
| 9 (\$100,000 or more) | 334 (15%) | 37 (6.3%) | 371 (13%) |
| (Missing) | 104 | 30 | 134 |
| Years of education, 50% (IQR) | 13 (12, 16) | 13 (12, 15) | 13 (12, 16) |
| Employment, n (%) |  |  |  |
| Employed | 943 (40%) | 72 (12%) | 1,015 (34%) |
| Not employed for wages | 159 (6.7%) | 33 (5.4%) | 192 (6.5%) |
| Retired | 722 (31%) | 308 (50%) | 1,030 (35%) |
| Unable to work (disabled) | 535 (23%) | 201 (33%) | 736 (25%) |
| (Missing) | 2 | 2 | 4 |
| Difficulty Paying Bills, n (%) |  |  |  |
| 1 (Very difficult) | 384 (16%) | 119 (20%) | 503 (17%) |
| 2 (Somewhat difficult) | 660 (28%) | 182 (30%) | 842 (29%) |
| 3 (Not very difficult) | 432 (18%) | 90 (15%) | 522 (18%) |
| 4 (Not at all difficult) | 865 (37%) | 219 (36%) | 1,084 (37%) |
| (Missing) | 20 | 6 | 26 |
| Number Admissions Past 12 months, 50% (IQR) | 0 (0, 2) | 2 (0, 3) | 1 (0, 2) |
| (Missing) | 24 | 7 | 31 |
| Elixhauser Score, 50% (IQR) | 7 (2, 13) | 15 (9, 22) | 9 (3, 16) |

**Abbreviations:** IQR – interquartile range; sTOFHLA – short test of functional health literacy; BHLS – Brief Health Literacy Screen; SNS – Subjective Numeracy Scale; ESSI – Enhancing Recovery in Coronary Heart Disease Emotional Social Support Instrument; MIDUS – Midlife in the United States; PHCS-2 – Perceived Health Competence Scale-2; BRCS – Brief Resilient Coping Scale; ARMS – Adherence to Refills and Medications Scale
**Note:** We display the median (25th, 75th percentile) for continuous variables, and frequency (percentage) for categorical variables. We used the Wilcoxon rank sum test for continuous variables and the Pearson Chi-square test for categorical variables.

During the follow-up period, 616 patients died. In descriptive analyses (Table 1), we observed that patients who died tended to be older, admitted for a heart failure exacerbation, had poorer health literacy, consumed less alcohol, exercised on fewer days, had lower income, were less likely to be employed, were more likely to be unable to work due to disability, had more hospital admissions in the last year, and had higher Elixhauser comorbidity scores.

Table 2 identifies patient-reported factors predictive of mortality in each of our analyses. The Base Model corresponds to the simple model that includes only the Demographic and Study Diagnosis domain variables. The Domain-Specific Models columns includes results from models that add each of the domains (separately) to the Base Model. Finally, the Full Model columns includes the final models with all domain variables included simultaneously. Each domain (Health literacy and Numeracy, Social support, Disease self-management, and Socioeconomic status) was found to be statistically significantly associated with mortality (p<0.01) when individually added to the Base Model (middle columns of Table 2).

**Table 2:**
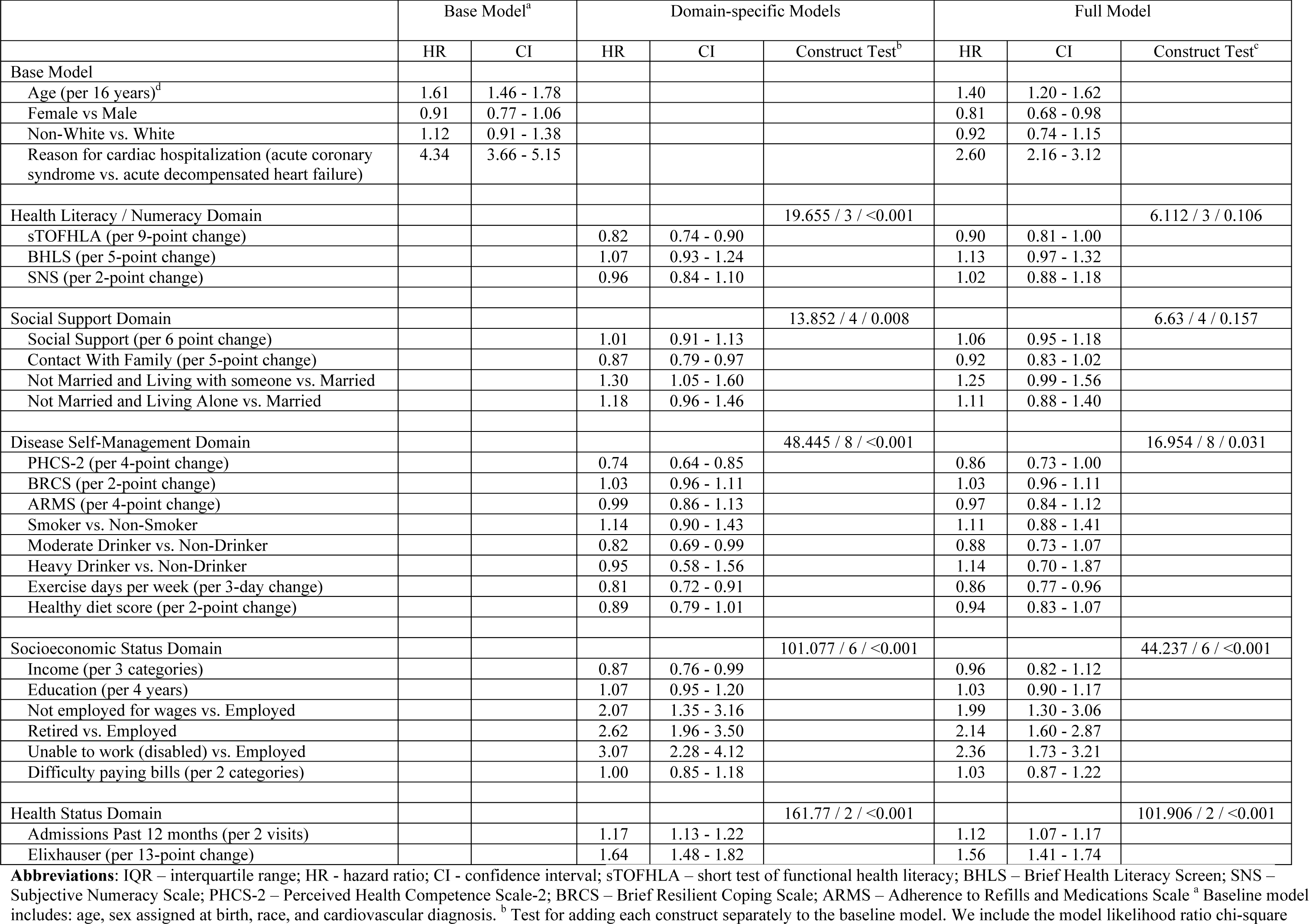
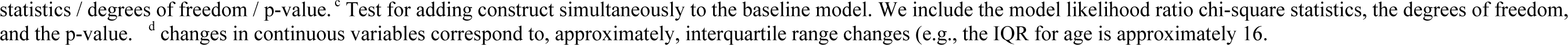
Cox regression analysis for risk of mortality after cardiovascular hospitalization.

|  | Base Model<sup>a</sup> |  | Domain-specific Models |  |  | Full Model |  |  |
| --- | --- | --- | --- | --- | --- | --- | --- | --- |
|  | HR | CI | HR | CI | Construct Test<sup>b</sup> | HR | CI | Construct Test<sup>c</sup> |
| Base Model |  |  |  |  |  |  |  |  |
| Age (per 16 years)<sup>d</sup> | 1.61 | 1.46 - 1.78 |  |  |  | 1.40 | 1.20 - 1.62 |  |
| Female vs Male | 0.91 | 0.77 - 1.06 |  |  |  | 0.81 | 0.68 - 0.98 |  |
| Non-White vs. White | 1.12 | 0.91 - 1.38 |  |  |  | 0.92 | 0.74 - 1.15 |  |
| Reason for cardiac hospitalization (acute coronary syndrome vs. acute decompensated heart failure) | 4.34 | 3.66 - 5.15 |  |  |  | 2.60 | 2.16 - 3.12 |  |
| Health Literacy / Numeracy Domain |  |  |  |  | 19.655 / 3 / <0.001 |  |  | 6.112 / 3 / 0.106 |
| sTOFHLA (per 9-point change) |  |  | 0.82 | 0.74 - 0.90 |  | 0.90 | 0.81 - 1.00 |  |
| BHLS (per 5-point change) |  |  | 1.07 | 0.93 - 1.24 |  | 1.13 | 0.97 - 1.32 |  |
| SNS (per 2-point change) |  |  | 0.96 | 0.84 - 1.10 |  | 1.02 | 0.88 - 1.18 |  |
| Social Support Domain |  |  |  |  | 13.852 / 4 / 0.008 |  |  | 6.63 / 4 / 0.157 |
| Social Support (per 6 point change) |  |  | 1.01 | 0.91 - 1.13 |  | 1.06 | 0.95 - 1.18 |  |
| Contact With Family (per 5-point change) |  |  | 0.87 | 0.79 - 0.97 |  | 0.92 | 0.83 - 1.02 |  |
| Not Married and Living with someone vs. Married |  |  | 1.30 | 1.05 - 1.60 |  | 1.25 | 0.99 - 1.56 |  |
| Not Married and Living Alone vs. Married |  |  | 1.18 | 0.96 - 1.46 |  | 1.11 | 0.88 - 1.40 |  |
| Disease Self-Management Domain |  |  |  |  | 48.445 / 8 / <0.001 |  |  | 16.954 / 8 / 0.031 |
| PHCS-2 (per 4-point change) |  |  | 0.74 | 0.64 - 0.85 |  | 0.86 | 0.73 - 1.00 |  |
| BRCS (per 2-point change) |  |  | 1.03 | 0.96 - 1.11 |  | 1.03 | 0.96 - 1.11 |  |
| ARMS (per 4-point change) |  |  | 0.99 | 0.86 - 1.13 |  | 0.97 | 0.84 - 1.12 |  |
| Smoker vs. Non-Smoker |  |  | 1.14 | 0.90 - 1.43 |  | 1.11 | 0.88 - 1.41 |  |
| Moderate Drinker vs. Non-Drinker |  |  | 0.82 | 0.69 - 0.99 |  | 0.88 | 0.73 - 1.07 |  |
| Heavy Drinker vs. Non-Drinker |  |  | 0.95 | 0.58 - 1.56 |  | 1.14 | 0.70 - 1.87 |  |
| Exercise days per week (per 3-day change) |  |  | 0.81 | 0.72 - 0.91 |  | 0.86 | 0.77 - 0.96 |  |
| Healthy diet score (per 2-point change) |  |  | 0.89 | 0.79 - 1.01 |  | 0.94 | 0.83 - 1.07 |  |
| Socioeconomic Status Domain |  |  |  |  | 101.077 / 6 / <0.001 |  |  | 44.237 / 6 / <0.001 |
| Income (per 3 categories) |  |  | 0.87 | 0.76 - 0.99 |  | 0.96 | 0.82 - 1.12 |  |
| Education (per 4 years) |  |  | 1.07 | 0.95 - 1.20 |  | 1.03 | 0.90 - 1.17 |  |
| Not employed for wages vs. Employed |  |  | 2.07 | 1.35 - 3.16 |  | 1.99 | 1.30 - 3.06 |  |
| Retired vs. Employed |  |  | 2.62 | 1.96 - 3.50 |  | 2.14 | 1.60 - 2.87 |  |
| Unable to work (disabled) vs. Employed |  |  | 3.07 | 2.28 - 4.12 |  | 2.36 | 1.73 - 3.21 |  |
| Difficulty paying bills (per 2 categories) |  |  | 1.00 | 0.85 - 1.18 |  | 1.03 | 0.87 - 1.22 |  |
| Health Status Domain |  |  |  |  | 161.77 / 2 / <0.001 |  |  | 101.906 / 2 / <0.001 |
| Admissions Past 12 months (per 2 visits) |  |  | 1.17 | 1.13 - 1.22 |  | 1.12 | 1.07 - 1.17 |  |
| Elixhauser (per 13-point change) |  |  | 1.64 | 1.48 - 1.82 |  | 1.56 | 1.41 - 1.74 |  |

**Abbreviations:** IQR – interquartile range; HR - hazard ratio; CI - confidence interval; sTOFHLA – short test of functional health literacy; BHLS – Brief Health Literacy Screen; SNS – Subjective Numeracy Scale; PHCS-2 – Perceived Health Competence Scale-2; BRCS – Brief Resilient Coping Scale; ARMS – Adherence to Refills and Medications Scale <sup>a</sup> Baseline model includes: age, sex assigned at birth, race, and cardiovascular diagnosis. <sup>b</sup> Test for adding each construct separately to the baseline model. We include the model likelihood ratio chi-square
statistics / degrees of freedom / p-value.<sup>c</sup> Test for adding construct simultaneously to the baseline model. We include the model likelihood ratio chi-square statistics, the degrees of freedom, and the p-value.<sup>d</sup> changes in continuous variables correspond to, approximately, interquartile range changes (e.g., the IQR for age is approximately 16).

The fully adjusted Cox proportional hazard models contained all domains simultaneously, as well as variables in the Base Model (the last columns of Table 2). Its estimated C-index was 0.79 consistent with moderate discrimination performance. The global, likelihood ratio chi-square statistic was 652.63, and on 27 degrees of freedom; this is consistent with minimal model overfitting (i.e., 4% shrinkage).^43^

The socioeconomic status and Health status domains were strongly associated with mortality (p<0.001) (Table 2). The Disease self-management domain was marginally associated with mortality (p=0.03) while the Health literacy/Numeracy and Social support domains were not. Within the Disease self-management and Socioeconomic Status domains, a few individual variables showed associations with mortality. A four point increase in the Perceived Health Competence Score was associated with a 14 percent decrease in hazard for mortality (Hazard Ratio (HR): 0.86, 95% Confidence Interval (CI): 0.73-1.00) in the Full Model. Similarly a three-day increase in the number of exercise days was associated with a 14 percent decrease in hazard for mortality (HR: 0.86, 95% CI: 0.77-0.96) in the Full Model. Employment status was strongly associated with mortality risk in the Full Model. Hazard ratios for those who were not employed, retired, and unable to work due to disability were 1.99, (95% CI 1.30-3.06), 2.14 (95% CI 1.60-2.87) and 2.36 (95% CI 1.73-3.21), respectively, when compared to those who were employed.

Figures 1a and 1b show relative contributions of the individual variables and domains to the information (i.e., variability in mortality explained) contained in the full model. We observed that reason for cardiac hospitalization and health status were the strongest contributors to model information. However, eight percent of the total variability explained in the model was due to the socioeconomic status construct. Further, of all variables, employment status contributed the third most information to the model.

**Figures 1a and 1b:**
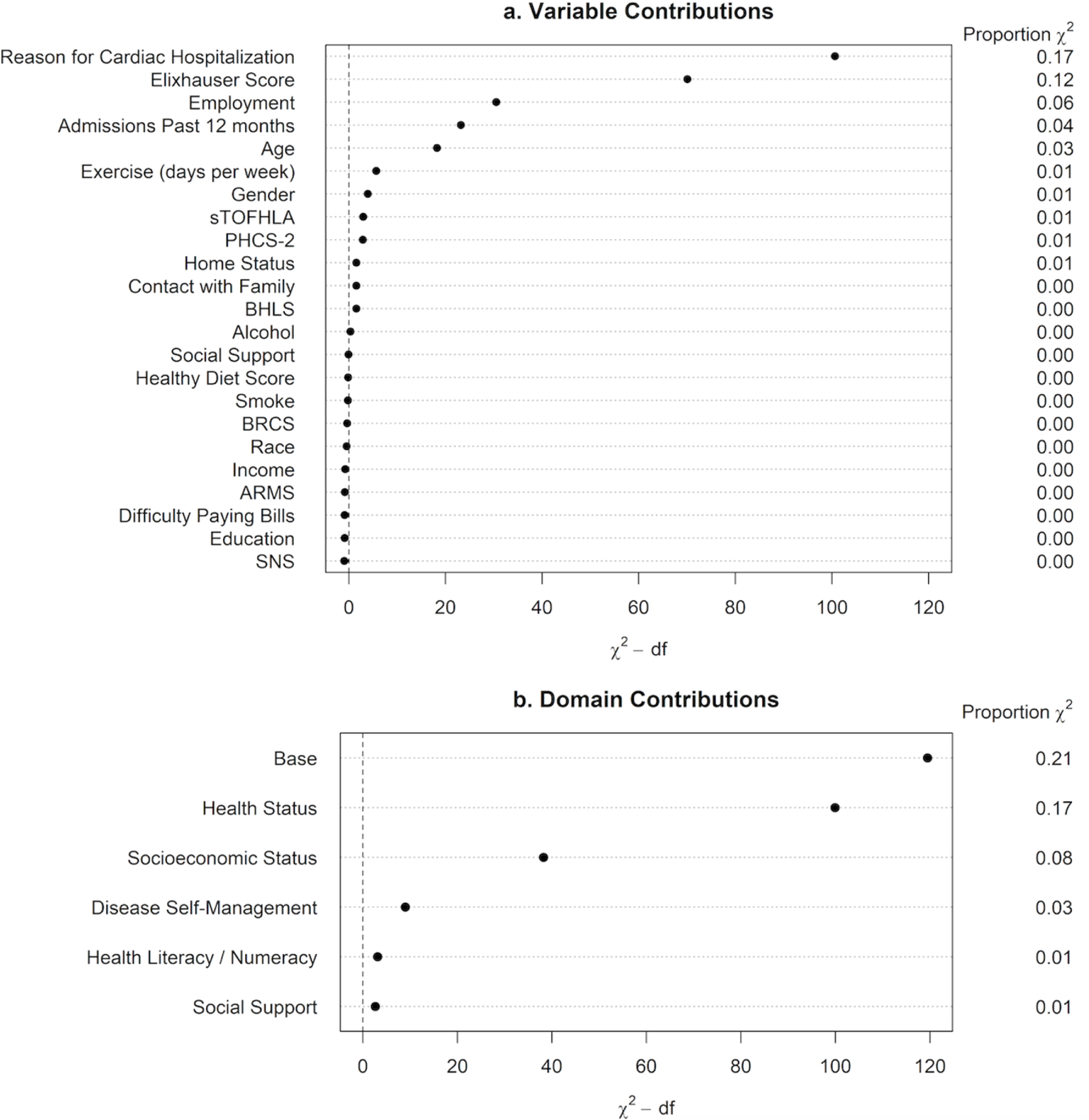
Relative explained variability plots. These figures display individual variable and domain contributions to the explained variability in the full model. We sorted the contribution by the likelihood ratio chi-square statistics corrected for the degrees of freedom and reported the relative explained variation (REV). REV was calculated using the Chi-square statistic associated with each variable or domain divided by the total Chi-square of the full model. It captures the proportion of information lost from the model if a single variable (Figure 1a) or if a domain of variables (Figure 1b) is removed from the full model.

## Discussion

To our knowledge, ours is the first large cohort that has assessed the mortality risk associated with a wide range of validated patient-reported measures in a high-risk cardiovascular population. Our results support that patient-reported measures of socioeconomic status and disease self-management may independently provide prognostic utility for adults hospitalized for cardiovascular events.

The addition of patient-reported measures to clinical data improves mortality risk prediction, though much of the literature thus far has focused on patients with malignancy. In an analysis of 3,240 men diagnosed with prostate cancer, the ten-year area under the received operating curve improved from 0.721 to 0.812 when patient-reported health measures were added.^44^ Cardiovascular hospitalizations are common and associated with increased morbidity and mortality, but few studies have determined the prognostic utility of patient-reported measures among adults hospitalized for these events to support successful transitions post-discharge.

In our analysis, employment status was the measure of socioeconomic status with the greatest prognostic significance. Patients were specifically asked whether they were unemployed due to disability. The significant, independent associations between being unemployed due to disability and mortality are striking. While our questionnaires did not ask patients to specify the type of disability they were experiencing, disability can often include physical function decline. Prior evidence demonstrates that physical function trajectory independently predicts cardiovascular and all-cause mortality, and we are currently investigating the prognostic utility measures of physical function (physical frailty and physical vulnerability) in populations with cardiovascular disease using validated measures.^45-46^ While our study design precludes our ability to identify the precise biological and behavioral mechanisms by which being disabled and/or unemployed contribute to mortality, our work further supports the utility of collecting these measures among adults hospitalized for cardiovascular events to identify those who may need additional post-discharge vocational or rehabilitative resources.

In our cohort, functional health literacy and exercise frequency were each individually predictive of mortality. We and others have previously demonstrated that functional health literacy is independently predictive of mortality among adults hospitalized for cardiovascular events, presumably through an inability to manage the complex medication prescriptions and dietary requirements associated with cardiovascular disease.^17,47^ Similarly, it has long been established that patient-reported exercise frequency is predictive of mortality among adults with increased risk for cardiovascular disease, though the precise amount of leisure time physical activity needed to confer a mortality benefit remains a source of ongoing investigation.^48^ However, our cohort added to these data by using a wide range of validated measures of disease self-management, followed patients for an extended period of time, and adjusted for numerous demographic and clinical characteristics.

Our analysis also included perceived health competence, an under-investigated construct that measures an individual’s sense of self-confidence in managing health conditions. In our cohort, perceived health competence was still marginally associated with long-term mortality even after adjustment for demographics, clinical comorbidities, and other patient-reported factors. Validity and reliability of the Perceived Health Competence Scale, which measures perceived health competence, has been established using adults living with a chronic illness.^20,32^ Given that the validated, abbreviated version applied in this cohort contains just two questions, incorporating these questions as part of a routine patient-facing assessment during hospital admission may be a novel, efficient way to identify those patients who need psychosocial support during transitions of care to reduce post-cardiac hospitalization mortality.

This study has several strengths. Few large cohorts exist that have included the type and range of validated patient-reported measures as has been included in our sample. In focusing on patient-reported measures, the study identifies factors that can alter the post-hospitalization trajectory of a group already at high risk for adverse outcomes. An additional strength is that long-term mortality data were obtained using thorough and rigorous methods, leading to robust outcome data.

Our study also has key limitations. Non-English speaking participants were excluded due to the fact that many of the patient-reported surveys used have only been validated in English. While this study was conducted at one academic medical center, the center has a very large, three-state catchement area. Those individuals who report unemployment due to disability may represent a group that also has a high comorbidity burden. More than likely, providers must aggressively treat these patients’ clinical comorbidities in addition to refer them to psychosocial or rehabilitative interventions to reduce mortality. Other limitations include that most adults in our cohort identified as white, this reducing the generalizability of our results across within other racial and ethnic groups. Finally, our patient-reported measures were only collected at baseline, limiting our ability to test for changes in these measures over time and their impact on outcomes.

## Conclusion

In a large cohort of adults hospitalized for a cardiovascular event, patient-reported measures of socioeconomic status, particularly employment, significantly and independently predicted post-discharge mortality at a median of 3.5 years after adjustment for demographic and clinical factors. Patient-reported measures of disease self-management; in particular exercise frequency and perceived health competence, were modestly associated with mortality after covariate adjustment. As collecting a large array of patient-reported measures takes time, focusing on these particular measures could be an efficient way to identify those patients who should be targeted for psychosocial and rehabilitative resources during the post-hospitalization period.

## Acknowledgements

We thank our study research analysts and project coordinators: Courtney Cawthon, Catherine Couey, Monika Rizk, Hannah Rosenberg, Daniel Lewis, Blake Hendrickson, Olivia Dozier, Vanessa Fuentes, Cardella Leak, Mary Lou Jacobsen, Catherine Evans, Joanna Lee, Emily Lucianno, and Erin Acord.

## Data Availability

All analytical data will be available upon request.

## References

1. Kaul P, Ezekowitz JA, Armstrong PW, Leung BK, Savu A, Welsh RC, et al. Incidence of heart failure and mortality after acute coronary syndromes. Am Heart J 2013 Mar;165(3):379–85.e2.

2. Johansson S, Rosengren A, Young K, Jennings E. Mortality and morbidity trends after the first year in survivors of acute myocardial infarction: a systematic review. BMC Cardiovascular Disorders 2017 17(53): 1–8.

3. Adler NE, Stead WW. Patients in context – EHR capture of social and behavioral determinants of health. N Engl J Med 2015 Feb 19;372(8):698–701.

4. Prather AA, Gottlieb LM, Giuse NB, et al. National Academy of Medicine Social and behavioral measures: associations with self-reported health. Am J Prev Med. 2017;53(4):449–456.

5. Roberts P, Deculus C, Garber L, Iivanainen A, Stentoft T, Winright K. Addressing social determinants of health: case studies from Epic’s Population Health Steering Board. Popul Health Manag. 2019;22(1):1–4.

6. Assess health-related social needs. The Joint Commission. Available at: https://www.jointcommission.org/our-priorities/health-care-equity/accreditation-standards-and-resource-center/assess-health-related-social-needs. Accessed March 29, 2023.

7. Doward LC, McKenna SP. Defining patient-reported outcomes. Value Health 2004 Sep-Oct;7 Suppl 1:S4-8.

8. Seow H, Tanuseputro P, Barbera L, et al. Development and validation of a prognostic survival model with patient-reported outcomes for patients with cancer. JAMA Netw Open. 2020;3(4):e201768.

9. Liao K, Wang T, Coomber-Moore J, Wong DC, Gomes F, Faivre-Funn F, Sperrin M, Yorke J, van der veer, Sabine. Prognostic value of patient-reported outcome measures (PROMs) in adults with non-small cell Lung Cancer: a scoping review. BMC Cancer 2022 Oct 19;22(1):1076.

10. Grossarth-Maticek R, Bastiaans J, Kanazir DT. Psychosocial factors as strong predictors of mortality from cancer, ischaemic heart disease and stroke: The Yugoslav prospective study. J Psychosom Res 1985 29(2):167–176.

11. Puterman E, Weiss J, Hives BA, Rehkopf DH. Predicting mortality from 57 economic, behavioral, social, and psychological factors. Proc Natl Acad Sci USA 2020 117(28):16,273-16,282.

12. Karraker A, Schoeni RF, Cornman JC. Psychological and cognitive determinants of mortality: evidence from a nationally representative sample followed over thirty-five years. Soc Sci Med 2015 Nov;144:69–78.

13. Everson-Rose SA, Lewis TT. Psychosocial factors and cardiovascular diseases. Annu Rev Public Health 2005. 26:469–500.

14. Ganna A, Ingelsson E. 5-year mortality predictors in 498,103 UK Biobank participants: a prospective population-based study. Lancet 2015 Aug 8;386(9993):533-540.

15. Patel CJ, Ioannidis JPA. Studying the elusive environment in large scale. JAMA 2014 Jun 4;311(21):2173–2174.

16. Korten AE, Jorm AF, Jiao Z, Letenneur L, Jacomb PA, Henderson AS, Christensen H, Rodgers B. Health, cognitive, and psychosocial factors as predictors of mortality in an elderly community sample. J Epidemiol Community Health 1999 Feb;53(2):83–88.

17. Mayberry LS, Schildcrout JS, Wallston KA, Goggins K, Mixon AS, Rothman RL, Kripalani S. Health literacy and 1-year mortality: mechanisms of association in adults hospitalized for cardiovascular disease. Mayo Clin Proc 2018 Dec;93(12):1728–1738.

18. Holt-Lunstad J, Smith TB, Layton JB. Social relationships and mortality risk: a meta-analytic review. PLoS Med 2010 Jul 27;7(7):e1000316.

19. Mokdad AH, Marks JS, Stroup DF, Gerberding JL. Actual causes of death in the United States, 2000. JAMA 2004 Mar 10;291(10):1,238-1,245.

20. Smith MS, Wallston KA, Smith CA. The development and validation of the Perceived Health Competence Scale. Health Educ Res 1995 Mar;10(1):51–64.

21. Meyers AG, Salanitro A, Wallston KA, Cawthon C, Vasilevskis EE, Goggins KM, et al. Determinants of health after hospital discharge: rationale and design of the Vanderbilt Inpatient Cohort Study (VICS). BMC Health Serv Res 2014 Jan 8;14:10.

22. Elixhauser A, Steiner C, Harris DR, Coffey RM: Comorbidity measures for use with administrative data. Med Care 1998, 36(1):8–27.

23. Assari S. General self-efficacy and mortality in the USA; racial differences. J Racial Ethn Health Disparities 2017 Aug;4(4):746–757.

24. Sarkar U, Ali S, Whooley MA. Self-efficacy as a marker of cardiac function and predictor of heart failure hospitalization and mortality in patients with stable coronary heart disease: Findings from the Heart and Soul Study. Health Psychol 2009 Mar 28(2): 166–173.

25. Self-management education: history, definition, outcomes, and mechanisms. Lorig KR, Holman H. Ann Behav Med 2003 Aug;26(1):1-7.

26. Nurss JR, Parker RM, Williams MV, Baker DW: Short test of functional health literacy in adults. Snow Camp, NC: Peppercorn Books and Press; 1998.

27. Chew LD, Bradley KA, Boyko EJ: Brief questions to identify patients with inadequate health literacy. Fam Med 2004, 36(8):588–594.

28. Fagerlin A, Zikmund-Fisher BJ, Ubel PA, Jankovic A, Derry HA, Smith DM: Measuring numeracy without a math test: development of the subjective numeracy scale. Med Decis Making 2007, 27(5):672–680.

29. Juster FT, Suzman R: An overview of the health and retirement survey. J Hum Resources 1995, 30(suppl):S7–S56.

30. Rossi AS: Social responsibility to family and community. In How healthy are we? A national study of well-being at midlife. Edited by Brim OG, Ryff CD, Kessler RC. Chicago: University of Chicago Press; 2004:550–585.

31. Mitchell PH, Powell L, Blumenthal J, Norten J, Ironson G, Pitula CR, Froelicher ES, Czajkowski S, Youngblood M, Huber M, et al: A short social support measure for patients recovering from myocardial infarction: the ENRICHD social support inventory. J Cardiopulm Rehabil 2003, 23(6):398–403.

32. Bachmann JM, Goggins KM, Nwosu SK, Schildcrout JS, Kripalani S, Wallston KA. Perceived health competence predicts health behavior and health-related quality of life in patients with cardiovascular disease. Patient Educ Couns 2016 Dec; 99(12):2071–2079.

33. Sinclair VG, Wallston KA: The development and psychometric evaluation of the brief resilient coping scale. Assessment 2004, 11(1):94–101.

34. Kripalani S, Risser J, Gatti M, Jacobson TA: Development and evaluation of the adherence to refills and medications scale (ARMS) among low literacy patients with chronic disease. Value Health 2009, 12(1):118–123.

35. Centers for Disease Control and Prevention: Behavioral risk factor surveillance system. Available at http://www.cdc.gov/brfss/index.htm. Accessed March 10, 2010.

36. Paxton AE, Strycker LA, Toobert DJ, Ammerman AS, Glasgow RE: Starting the conversation performance of a brief dietary assessment and intervention tool for health professionals. Am J Prev Med 2011, 40(1):67– 71.

37. Sallis R: Developing healthcare systems to support exercise: exercise as the fifth vital sign. Br J Sports Med 2011, 45:473–474.

38. Chen J, Rathore SS, Radford MJ, Wang Y, Krumholz HM. Racial differences in the use of cardiac catheterization after acute myocardial infarction. N Engl J Med 2001;344:1443–1449.

39. Maynard C. The incompleteness of the social security death master file. JAMA Cardiol 2019;4(8):831.

40. R Core Team (2022). R: A Language and Environment for Statistical Computing. R Foundation for Statistical Computing, Vienna, Austria. https://www.R-project.org/.

41. Harrell Jr FE (2021). rms: Regression Modeling Strategies. R package version 6.2–0, https://CRAN.R-project.org/package=rms.

42. Therneau T (2021). A Package for Survival Analysis in R. R package version 3.2–13, https://CRAN.R-project.org/package=survival.

43. Van Houwelingen JC, Le Cessie S. Predictive value of statistical models. Statist Med. 1990;9(11):1303-1325.

44. Bachmann JM, Posch DR, Hickson GB, Pinson CW, Kripalani S, Dittus RS, Stead WW. Developing an implementation strategy for systematic measurement of patient-reported outcomes at an academic health center. J Health Manag 2020 Jan-Feb;65(1):15-28.

45. Tan H-J, Zhou X, Sprattle BN, McMahon S, Nielsen ME, Lung J, Harris AHS, Smith AB, Basch E. Patient-reported vs claims-based measures of health for modeling life expectancy in men with prostate cancer. J Urol 2021 Feb;205(2):434–440.

46. Fried LP, Kronmal RA, Newman AB, Bild DE, Mittelmark MB, Polak JF, Robbins JA, Gardin JM. Risk factors for 5-year mortality in older adults: the Cardiovascular Health Study. JAMA 1998;279(8):585–592.

47. Hu Z, Zheng B, Kaminga AC, Zhou F, Xu H. Association between functional limitations and incident cardiovascular disease and all-cause mortality among the middle-aged and older adults in China: a population-based prospective cohort study. Front Public Health 2022; 10:751985.

48. Moser DK, Robinson S, Biddle MJ, Pelter MM, Nesbitt T, Southard J, Cooper L, Dracup K. Health literacy predicts morbidity and mortality in rural patients with heart failure. J Card Fail 2015 Aug; 21(8):612–618.

49. Lee DH, Rezende LFM, Joh HK, Keum N, Ferrari G, Reylopez JP, Rimm EB, Tabung FK, Giovannucci EL. Long-term leisure-time physical activity intensity and all-cause and cause-specific mortality: a prospective cohort of US adults. Circ 2022;146:523-53.

